# What factors influence symptom reporting during an emerging infectious disease outbreak? A rapid review of the evidence

**DOI:** 10.1101/2020.07.23.20159897

**Authors:** Patrice Carter, Odette Megnin-Viggars, G James Rubin

## Abstract

**Introduction:** During any emerging infectious disease outbreak, people with symptoms of the illness are often asked to report their symptoms to the health service in a timely manner, to facilitate contact tracing. Numerous factors may influence an individual’s willingness to report these symptoms. Understanding these factors has become urgent during the COVID-19 pandemic.

**Objective:** To determine which factors influence symptom reporting during an emerging infectious disease outbreak.

**Methods:** We conducted a rapid review of the evidence. We included papers based on primary research; published in a peer-reviewed journal; written in English; included factors associated with symptom reporting or accessing healthcare; and related to a major public health incident involving an infectious disease outbreak.

**Main results:** Five themes were identified as facilitators of symptom reporting or accessing healthcare: accurate and informative communication; symptom severity; concern about exposure; ease of access; and relationship with the healthcare provider. Seven themes were identified as barriers of symptom reporting or accessing healthcare: lack of knowledge; fear; stigmatization; invasion of privacy; low concern about symptoms; economics; and practicalities of attending a healthcare facility.

**Discussion & Conclusion:** If contract tracing services are to be effective, members of the public need to have the capability, opportunity and motivation to use them. The themes identified should be used to evaluate the information provided to the public about such a service, the routes of access, and the underlying policies relating to the service, in order to ensure that as many people as possible with relevant symptoms will make contact.

## Introduction

Governments across the world are using contact tracing services in order to provide a route out of lockdown from COVID-19. In the UK, the Government has launched a “Test, Trace and Isolate” system. This requires people who develop a new, continuous cough, a fever, or a loss of their sense of taste or smell (the ‘index case’) to follow a series of actions. First, they must book a test through a drive-through facility or have a test delivered to their home. Second, they (and any household members) must remain at home until they receive their test result. Third, for those who test positive, further isolation is advised and a process of tracing the person’s close contacts begins. Contacts are asked to quarantine themselves, and to request a test if they too, develop symptoms.

Every step of this system relies on people adhering to advice. To date, most attention has focused on the challenges involved in helping index cases or their contacts adhere to advice to remain in isolation or quarantine for the recommended period of time (1). However, challenges exist at every step. In this paper, we focus on the very first steps in the patient pathway: recognizing that one has relevant symptoms and reporting these to a health service (or in the case of the UK, deciding to request a test).

This process can itself be broken down into stages. Symptom perception is determined partly by physiological factors. These might include infection with COVID-19, but physical signs of illness can also be triggered by other medical factors or environmental factors. Symptom perception is influenced by multiple psychological and contextual factors, being more likely to occur among people who expect to experience a given symptom, or who are worried or anxious, and are hence monitoring themselves for symptoms (2, 3).

Seeking help, once a symptom has been perceived is then also determined by a range of factors. The person’s attribution of their symptoms plays a key role; most symptoms experienced in everyday life are appraised as being transient and benign, and are not brought to the attention of the health services. This appraisal is partly based on the severity, suddenness and duration of the symptom. Minor, gradually evolving and transient symptoms are less likely to be interpreted as indicating something troubling (4). However, awareness of specific health risks is also important. For example, not recognizing a symptom as a possible warning sign is a key factor underlying delayed help seeking for cancer (5, 6) and rheumatoid arthritis (4, 7, 8).

Regardless of the cause of an individual’s symptoms, once the person has decided they have a problem, the decision to seek help can be complex and result in delay. For example, an analysis in England found that of the 1732 people who described having what the authors termed an ‘alarm’ symptom for cancer over the past three months, 28.4% had not yet consulted a doctor about an unexplained lump, 30.4% had not sought help for unexplained pain, and 45.8% had not sought help for unexplained bleeding (9). Important factors influencing the decision to seek help include the fear of being seen as ‘a time-waster’ (4, 5, 10), competing demands on one’s time (5, 10), worries about the impact or efficacy of any intervention that might be offered (4, 5), pressure from friends and family (11, 12) and perceptions that one is complying with official advice or using a service appropriately (13-15).

We are not aware of any currently published systematic reviews which explore factors associated with facilitating or deterring people from reporting symptoms during major health incidents involving an infectious disease outbreak. In this paper, we report a rapid review of the literature, to determine what factors influence symptom reporting during such an incident.

## Methods

Following the PRISMA guidelines (16) we developed a protocol for a rapid review to determine what factors influence symptom reporting during major health incidents.

### Search strategy

We developed a search strategy which included medical subject headings and free text terms. Key words for the search included terms for epidemics/pandemics (including coronavirus, avian influenza, Ebola, Middle East respiratory syndrome, severe acute respiratory syndrome, swine flu) and terms characterizing symptom reporting (including help-seeking behaviour, symptom reporting, stigma, requesting/asking for a test). Three electronic databases (Medline, PsycINFO, ProQuest) were searched from inception to June 2020, the full list of search terms can be found in Appendix A.

### Selection criteria

For studies to be included in this review, they had to: report on primary research; be published in a peer-reviewed journal; be written in English; include factors associated with reporting symptoms to a healthcare unit, or accessing healthcare (i.e. facilitators, barriers, or features associated with the reporting of symptoms or accessing of healthcare) and include participants with experience of a major health incident, which had to be viral, contagious and not sexually transmitted. Citations from each database search were downloaded into EndNote and duplicates removed. Titles and abstracts of identified studies were screened by two reviewers for inclusion against criteria, until a good inter-rater reliability had been observed (percentage agreement =>90%). Initially 10% of references were double-screened, and as inter-rater agreement was good (98%), the remaining references were screened by one reviewer. All primary-level studies included after the first screen of citations were acquired in full and re-evaluated for eligibility (see Appendix B for studies that were excluded at full-text review).

### Data extraction and synthesis

Data extraction and synthesis was conducted independently by two reviewers, and discrepancies or difficulties with coding were resolved through discussion. Quality of each included study was assessed using the CASP (Critical Appraisals Skills Programme) Qualitative checklist for qualitative studies (17) or the BMJ Critical appraisal checklist for survey studies (18). A quality rating was assigned to each study, where ++ indicates that most (≥75%) or all of the checklist criteria have been met, + indicates that the checklist criteria have been partially met (≥50%-75%), and – indicates that the majority of checklist criteria have not been met (<50%). Thematic synthesis was used to inductively identify emerging themes in qualitative findings across studies. Themes from individual studies were integrated into a wider context, with overarching categories of themes and subthemes being developed. Themes were derived from the data presented within the included studies, emerging themes were placed into a thematic map representing the relationship between the overarching categories, themes and subthemes. Narrative synthesis was used to combine and summarise the findings from survey studies, this was embedded within the context of the qualitative findings to provide an overall narrative of the data.

## Results

The systematic search of electronic databases generated a total of 2107 references, and two additional references were identified through hand searching. Of these 2109 references, 58 were reviewed in full-text. Sixteen studies met the eligibility criteria (see Figure 1 for flow chart of study search and selection process).

**Figure 1:**
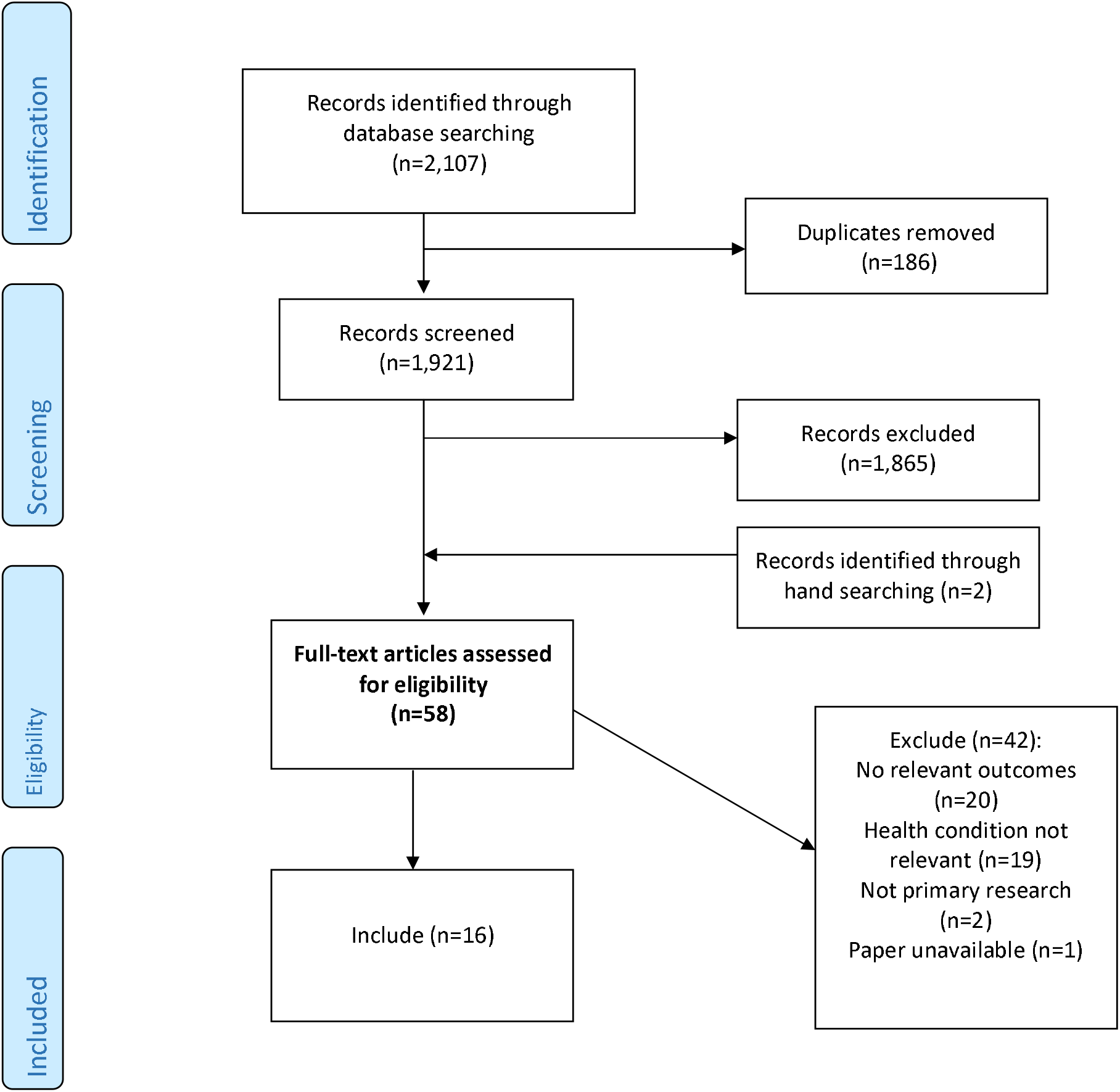
PRISMA flow chart.

### Included studies

The characteristics of the included studies are summarised in Table 1. Half the studies used qualitative methods (11, 19-25) and half used quantitative surveys (26-33).

**Table 1:** Characteristics of included studies.

| Author and country | Study design | Infectious disease | Participants | Study aim | Sample size | Percentage female | Percentage BAME (non-white) | Included Themes | Study quality* |
| --- | --- | --- | --- | --- | --- | --- | --- | --- | --- |
| Aghaizu 2011 (19)<br>England, Germany, Greece & Hungary | Qualitative study; focus groups | Any infectious disease | Hospital based healthcare workers | To determine Healthcare workers' attitudes to monitoring their sickness, absence and reporting infectious disease symptoms | 49 | NR | NR | Facilitators of symptom reporting<br>Accurate & Informative communication<br>Symptom severity<br>Barriers to symptom reporting<br>Fear<br>Lack of knowledge<br>Invasion of privacy<br>Economics<br>Stigmatisation | + |
| Biggerstaff 2012 (26)<br>US | Quantitative study; telephone survey | H1N1 influenza like illness (ILI) | General adult population reporting ILI symptoms | To characterise people reporting ILI and to evaluate trends in healthcare seeking behaviours, clinical diagnosis and treatment of pH1N1 | 216, 431 | 62 | 22 | Associated features of reporting symptoms<br>Demographics | + |
| Brooks-Pollock (27)<br>England | Quantitative study; online survey | H1N1 influenza like illness (ILI) | General adult population reporting ILI symptoms | To quantify healthcare seeking behaviour during the 2009 H1N1v epidemic | 1,522 | NR | NR | Associated features of reporting symptoms<br>Demographics | + |
| Carter 2017 (20)<br>Sierra Leone | Qualitative study; face to face interview | Ebola | Adults living in Ebola-affected communities | To determine barriers and enablers of treatment seeking behaviour for Ebola | 35 | NR | NR | Facilitators of symptom reporting<br>Ease of access<br>Relationship with healthcare provider<br>Barriers to symptom reporting<br>Fear<br>Lack of knowledge | + |
| Carter 2017a (21)<br>Sierra Leone | Qualitative study; face to face interviews, focus groups and | Ebola | Adults living in Ebola-affected communities | To determine barriers and enablers of treatment seeking behaviour for Ebola | 350 | NR | NR | Facilitators of symptom reporting<br>Accurate & Informative communication<br>Relationship with healthcare provider | + |
|  | questionnaires |  |  |  |  |  |  | Barriers to symptom reporting<br>Low concern about symptoms |  |
| Kjelso 2016 (28)<br>Denmark | Quantitative study; online data from "Influmeter" website | Influenza like illness (ILI) | General adult population reporting ILI symptoms | To describe the participants representatively using a new online system (as compared to the Danish population) and to compare the signal detection to existing surveillance systems | 1,089 | 67 | NR | Associated features of those reporting symptoms<br>Demographics | - |
| Kreslake 2016 (29) | Quantitative study; healthcare | Avian influenza (HPAI) | General adult population reporting ILI | To determine the use of healthcare facilities, perceptions of | 2,520 | NR | NR | Facilitators of symptom reporting<br>Symptom severity | - |
| Indonesia | utilisation survey and face to face survey | H5N1) | symptoms (survey data). Interviews conducted in those with and without symptoms | healthcare services and intentions to seek treatment in hypothetical scenarios |  |  |  | Barriers to symptom reporting<br>Low concern about symptoms<br>Economics<br>Practicalities of attending healthcare facility<br>Stigmatisation<br>Associated features of those seeking healthcare<br>Demographics |  |
| Lohiniva 2011 (22)<br>Egypt | Qualitative study; focus groups and semi-structured interviews | Acute respiratory illness (ARI) | Household caregiver (defined as the primary caretaker of children or | To explore the classification of ARI, to understand modes of transmission, symptoms of severity, and to explore health | 18 focus groups and 20 interviews | 100 | NR | Facilitators of symptom reporting<br>Ease of access<br>Relationship with healthcare provider | + |
|  |  |  | other household members during ARI episodes | seeking behaviours and home management practices |  |  |  | Barriers to symptom reporting<br>Low concern about symptoms<br>Stigmatisation |  |
| Manabe 2012 (30)<br>Vietnam | Quantitative study; face to face interviews | Avian influenza (H5H1) | General population of those living in -affected and non-affected communities | To assess knowledge, attitude, practice and emotional response to H5N1 | 543 | 68 | NR | Facilitators of symptom reporting<br>Symptom severity<br>Accurate & informative communication | + |
| McLean 2018 (23)<br>Liberia | Qualitative study; semi-structured interviews | Ebola | Individuals from households who reported being sick | To explore experiences of illness, impact of disease and mortality | 226 | 55 | NR | Barriers to symptom reporting<br>Practicalities of attending healthcare facility<br>Economics<br>Fear<br>Stigmatisation<br>Low concern about symptoms | - |
| Meng 2016 (31)<br>Hong Kong | Quantitative study; telephone surveys | Influenza like illness (ILI) | General adult population | To compare healthcare seeking behaviours and self-medication of ILI patients between summer and winter influenza epidemics | 1,040 | 62 | NR | Facilitators of symptom reporting<br>Accurate & informative communication<br>Associated features of those reporting symptoms<br>Demographics | + |
| Rubin 2010 (11)<br>UK | Qualitative study; semi-structured interviews | Influenza A (H1N1) | General adult population | To understand why people contacted the health service about swine flu (H1N1), and to understand patients' motivations for seeking advice | 5,419 | 58 | 27 | Facilitators of symptom reporting<br>Symptom severity<br>Concern about exposure | ++ |
| Rubin 2010a (32) | Quantitative study; telephone survey | Influenza A (H1N1) | General adult population | To determine associations between worry about catching swine flu (H1N1) and | 33 | 59 | 7 | Barriers to symptom reporting<br>Low concern about symptoms<br>Concern about exposure | + |
| UK |  |  |  | level of media coverage, and the role of media coverage and advertising in predicting accessing healthcare |  |  |  | Associated features of those reporting symptoms<br>Demographics |  |
| Soyemi 2009 (24)<br>US | Qualitative study; telephone interviews | Influenza (pH1N1) | People from Illinois who had been hospitalised for pH1N1 | To identify individual level factors relating to health seeking behaviour | 2,824 data surveillance records and 33 interviews | 53 (surveillance records) and 42 (interviews) | 55 | Barriers to symptom reporting<br>Practicalities of attending healthcare facility<br>Economics<br>Fear<br>Stigmatisation<br>Low concern about symptoms<br>Associated features of those reporting symptoms<br>Demographics | + |
| Tate 2017 (33)<br>US | Quantitative study; electronic or mailed survey | Ebola | People from New York City who had been part of the active monitoring scheme for Ebola | To evaluate frequency of reporting false data during active Ebola monitoring, factors associated with reporting false data, and the psychosocial impact of preferences during the Ebola active monitoring | 393 | 41 | NR | Barriers to symptom reporting<br>Fear<br>Low concern about symptoms<br>Lack of knowledge | + |
| Yamanis 2016 (25)<br>Sierra Leone | Qualitative study; semi-structured interviews | Ebola | Adults living in Ebola-affected communities | To explore perceptions and intentions of those using the Ebola response system | 30 | 47 | NR | Facilitators of symptom reporting<br>Ease of access<br>Barriers to symptom reporting<br>Fear<br>Low concern about symptoms | ++ |
|  |  |  |  |  |  |  |  | o Lack of knowledge<br>o Economics |  |

\*Study quality: Assessed using relevant tool; CASP for qualitative studies and BMJ survey checklist for quantitative surveys: Scores = ++ most of checklist criteria met; + some of checklist criteria met; - insufficient checklist criteria met.

The studies were conducted across 12 countries and included people with experience of the 2009 and 2010 H1N1 influenza pandemic (11, 24, 26, 27, 32), Ebola (20, 21, 23, 25, 33), avian influenza (29, 30), acute respiratory illness (22), influenza-like illness (28, 31) and any infectious disease (19).

### Identified themes

Five themes were identified as facilitators of symptom reporting or accessing healthcare: accurate and informative communication; symptom severity; concern about exposure; ease of access; relationship with healthcare provider. Seven themes were identified as barriers of symptom reporting or accessing healthcare: lack of knowledge; fear; stigmatization; invasion of privacy; low concern about symptoms; economics; practicalities of attending healthcare facility. See Figure 2 and 3 for theme maps for facilitators and barriers of symptom reporting. These themes (and subthemes) are elaborated below. The synthesis of quantitative survey data also highlighted a number of demographic features associated with those reporting symptoms and those seeking healthcare, summarised below.

**Figure 2:**
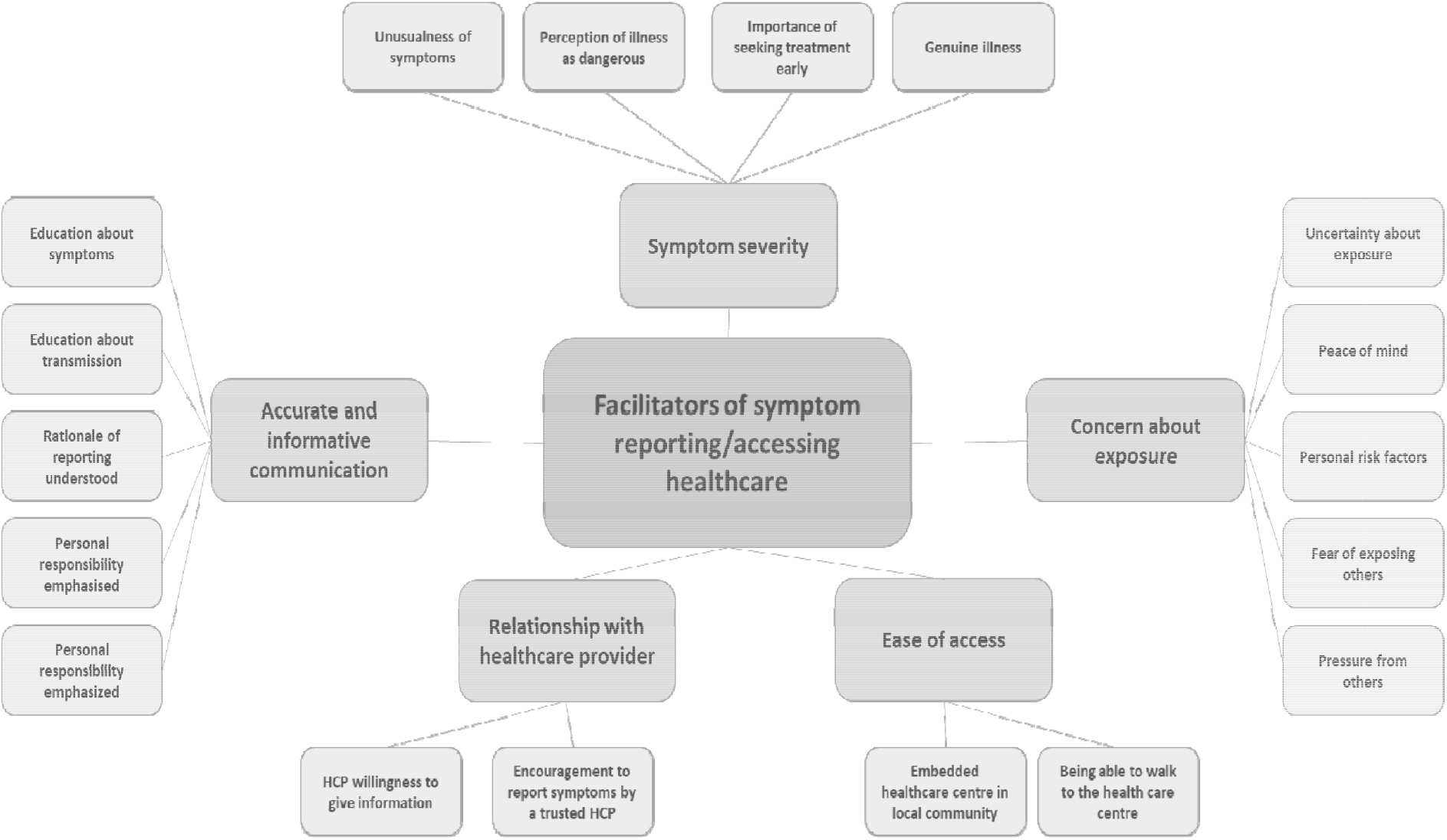
Facilitators of symptom reporting theme map.

**Figure 3:**
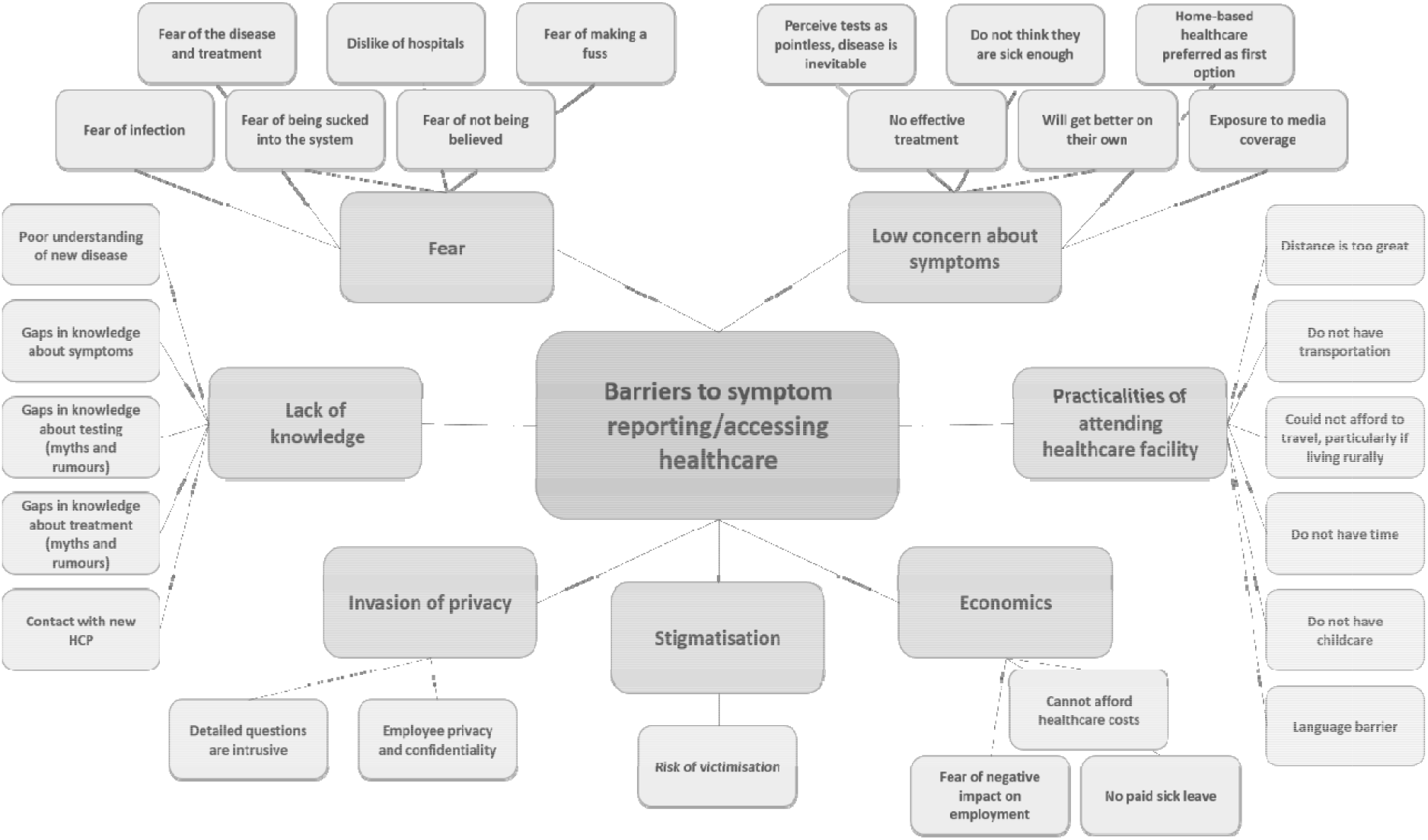
Barriers of symptom reporting theme map.

## Facilitators of symptom reporting/accessing healthcare

### Accurate and informative communication

Participants found that education about symptoms (19, 21, 30, 31) and about routes of transmission enabled them to feel confident about reporting symptoms and seeking help. Participants emphasized that they would be happier to report symptoms if they understood the rationale behind reporting, and if their personal responsibility was emphasised (19):

*“I think most people would be happy if they knew there was a risk, if you could tell them why you’re asking the questions*.*”* ((19), pg. 9)

Participants also highlighted that having confidence that the reporting system was effective and efficient would facilitate reporting (19).

### Symptom severity

Participants reported that the unusualness of symptoms, particularly in terms of severity and duration, were powerful prompts to access healthcare (11, 29)

*“I’ve had a cold and I’ve had the flu and it only lasted about 2 days. But I had this for about a week and a half. It’s the first I’ve had a cold that long and I was just thinking the worst”* ((11) pg. 3)

Early access to healthcare was also facilitated when people perceived the illness as dangerous (29, 30). Participants were more motivated to seek treatment earlier when the importance of early response was emphasised, and where exposure to survivors built hope about recovery (21, 29). Participants expressed the view that if they were confident that they were experiencing a genuine illness they would feel comfortable about disclosing symptoms (19).

### Concern about exposure

Participants were motivated to seek advice if they were concerned or uncertain about exposure (11). Another related motivator was a desire to gain peace of mind (11). Furthermore, those with personal risk factors (including age and underlying health conditions) were particularly compelled to contact healthcare services (11). Social pressures also facilitated help-seeking behaviour, both in terms of fears about exposing others to the disease, and pressure from others to report symptoms (11).

### Ease of access

The physical proximity of healthcare facilities and the importance that these were local and community-based was highlighted as facilitators to accessing healthcare as the people working in those facilities would be more likely to be known and trusted (20, 25). Being able to walk to facilities also resolved transportation issues from a practical perspective, and provided an emotional facilitating factor as many participants described an intense fear of ambulances (20, 22)

### Relationship with healthcare provider

Participants were more likely to seek help if they were encouraged to do so by familiar and trusted healthcare professionals (20). Help-seeking behaviour was also facilitated through healthcare professional’s willingness to discuss symptoms and provide information (22). Conversely, having to contact a new and unknown healthcare provider was highlighted as a barrier that delayed the decision to seek healthcare (24).

### Barriers of symptom reporting/accessing healthcare

#### Lack of knowledge

Participants described a poor understanding of the characteristics of a new disease and how this could act as a barrier to help-seeking behaviour (19, 25). More specifically, participants talked about gaps in knowledge about symptoms (33) gaps in knowledge about testing (20, 25) and gaps in knowledge about treatment (20, 25) and how these gaps could be filled with rumours and myths that could make people even more fearful about accessing healthcare:

*“There were rumors that the needle to test you actually is poison and the test kills you*.*”* ((20), pg. 68)

### Fear

Fear emerged as a recurrent barrier to symptom reporting, this included: fear of the what may happen if a person caught the disease, fear of receiving treatment, and of the treatment system (20); fear of being infected in a healthcare setting (23); fears of being sucked into the system that arose when people witnessed sick people being taken away and never returning (20, 25); fear of the hospital or clinic environment (24, 25, 33); fear of the reaction they may receive from the healthcare worker, i.e. of not being believed (19) or of being seen to make a fuss (33).

### Stigmatisation

Participants anticipated stigmatization and the desire to avoid this created a barrier to reporting symptoms (19, 22, 23). People worried about experiencing stigma in the workplace, in the wider community, and were concerned about social stigma that may be attached to family members of those who had developed severe illness.

### Invasion of privacy

Participants were wary that detailed questioning about symptoms could be experienced as an invasion of privacy and create or increase feelings of anxiety and distrust (19). The need for the protection of employee privacy and confidentiality was also raised, in the context of healthcare workers reporting symptoms (19).

### Low concern about symptoms

Juxtaposed against the finding that concern about exposure could act as a facilitator to symptom reporting, others experienced low concern about symptoms and this acted as a barrier to help-seeking. For some people, this took the form of a fatalistic approach, with tests being perceived as pointless and disease inevitable (21), or the lack of an effective treatment delaying the decision to seek healthcare (24). Others just did not feel sufficiently ill to report symptoms or access healthcare (25, 29, 33):

*“Because based on what the medical people say, if it is high fever, then you will say this has gone beyond limit. Because there are times when we get malaria and common cold before this Ebola. So you cannot just start feeling feverish and you say you have to call 117*.*”* ((25) pg. 7)

Some participants believed that there was no need to access healthcare as they would get better on their own (22, 29), or preferred to try home-based healthcare as a first-line treatment option (23-25).

One study found that exposure to media coverage reduced worry and decreased healthcare service use. (32)

### Economics

Economic barriers to accessing healthcare were also experienced, because participants could not afford healthcare (23-25), or were worried about a negative impact on their employment or had no paid sick leave (24).

### Practicalities of attending a healthcare facility

For some people the healthcare facility was too far away to travel to (29), and a lack of transportation (23, 24, 29), or not being able to afford to travel (29) imposed practical barriers to accessing healthcare, particularly for those living in rural communities. Other practical barriers to help-seeking behaviour included: not having time to visit a healthcare facility (29); lacking childcare (29); and a language or communication barrier (24).

### Associated features of those reporting symptoms

Three studies found that women were more likely to report symptoms than men (26, 28, 31), we cannot however, determine if this is because women were more likely to contract the illness. Two studies reported that older people (aged 65 years or over) were less likely to report symptoms than those who could be categorized as younger or middle-aged, although these age bands were large, covering 18 to 49 year olds (26) or 25-64 years (28). One study found that white participants were more likely to report symptoms than black participants (26). However, the relationship of ethnicity and reporting was more complex than all black and minority ethnic (BAME) groups being less likely to report symptoms, as the same study (26) found that American Indian/Alaska Native participants were more likely to report symptoms than white adults. Finally, two studies found that people with higher levels of education were more likely to report symptoms than those who had spent less time in education.

### Associated features of those seeking healthcare

Two studies found that women were more likely to seek medical care than men (26, 31). In an opposite pattern to the reporting of symptoms, one study reported that those with primary level education or below were more likely to seek medical care than those with tertiary education or above (31). One study found that people from ethnic minority groups were more likely to access healthcare (32). Two studies reported that those with underlying health conditions (31, 32) were more likely to have sought medical care, and one of those studies also found that people living in larger households (Rubin 2010b) were more likely to access healthcare. Again, within this review we cannot determine if greater levels of seeking healthcare by these groups is due to an increased likelihood that they have contracted the illness. Finally, mixed results were reported across studies on the association between age and healthcare seeking behaviour, with two studies finding that older adults (aged at least 60 or 65 years) were more likely to seek healthcare (26, 31), and another study in which participants indicated that they felt too old to seek care (29). Another two studies found no direct association between age and help-seeking behaviour, but found relationships with other variables. One study reported a relationship between age and gender with males under the age of 25 being more likely to seek medical attention than women under the age of 25 (27). Another study reported a relationship between age and ethnicity, as for BAME respondents the longest duration between onset of symptoms and seeking initial treatment was found for an older age group, while for white respondents the longest duration was reported by children and young people (24).

## Discussion

### Principal finding

The evidence within this review shows a number of distinct factors which can act as either facilitators or barriers to people reporting symptoms to, or seeking care from, the health services during an emerging infectious disease outbreak. These include fear or concern about exposure, knowledge & information, level of concern over symptoms, and practical considerations. Additional factors are related to invasion of privacy and stigmatisation. A number of demographic factors should also be considered, including sex, age, ethnicity and education.

### Interpretation of results

Our thematic analysis identified four main themes which facilitate people either reporting symptoms or accessing health care facilities: symptom severity, concern about exposure, accurate and informative communication, relationship with health care provider and ease of access. For example when information is accurate, and communicated effectively people understand why they need to report symptoms, and feel enabled to do so (19), whereas when people have a poor understanding, either about testing, the disease, or disease symptoms they appear reluctant to visit a healthcare facility or report symptoms (20, 25). Public health officials need to ensure people receive appropriate information, working with the media, who during major health crisis play an important role (34). Calls for good communication are, of course, nothing new in the field of emergency response. However, we make no apologies for raising it once more. Without sustained effort on the part of official communicators, knowledge among the public can be surprisingly low. For example, five months into the COVID-19 pandemic only 59% of the UK population knew what symptoms they were being asked to be alert to (35). Additionally, the role of healthcare professionals is important, the data showed that people were more positive when the healthcare provider seemed willing to give them information and talk to them (22).

Accurate and informative communication links closely to the identified themes, “symptom severity” (a facilitator), and the theme “low concern about symptoms” (a barrier). If people are not sufficiently educated about relevant symptoms they may not feel it is appropriate to access healthcare, or report their symptoms; therefore, clear advice on severity and nature of symptoms is needed. These data are consistent with other studies which have shown that when people think their symptoms are minor or unimportant, they are less likely to report them (4). When people believed they had a genuine illness, and that symptoms were severe or serious, they would report them (19). These findings are supported by studies which show the greater the perceived perception of illness severity, the more likely they are to engage in precautionary behaviours (36), for example contacting the healthcare service. While in daily life, such attitudes may be appropriate, during the COVID-19 outbreak, this represents a substantial problem for testing and contact tracing services. Given that people are infectious even if asymptomatic, it is essential to encourage people to report even mild symptoms without delay. A focus on this in messaging will be required, as our data suggest that this runs contrary to most people’s natural inclinations.

Anxiety about exposure also increased people’s motivations to seek advice or report symptoms. When people were concerned that their individual situation increased their susceptibility to a disease, they were more likely to engage with the healthcare system. In contrast, fear about what might happen next once ‘within the system’ was identified consistently across studies, where it acted as a barrier to reporting symptoms. These findings suggest that it is important to reduce fear about being infected by visiting a healthcare setting, or about receiving treatment, but instilling a sufficiently realistic degree of concern about the illness that motivates people to engage with the system.

Practical issues were regularly reported in the evidence. Having a health care facility embedded in the local community, and within walking distance was a facilitator for people reporting symptoms. However, when facilities were deemed too far to get to, too expensive to travel to, or too difficult to travel to, this was seen as a barrier. The cost of accessing the healthcare system was also a consideration for many. Indirect economic factors were also important. People reported difficulties with getting time of work and not having paid sick leave. During an emerging infectious disease outbreak, tackling economic barriers to seeking testing should be an essential component of any fully functioning public health response.

Evidence on the associated features for those seeking healthcare and those reporting symptoms showed clear differences between women and men, different age groups, and people from different ethnic backgrounds. Studies showed that women consistently reported symptoms more often, and were more likely to seek healthcare than men; highlighting that more needs to be done to engage the male population. However, the data were not consistent in relation to age or ethnicity.

### Strengths & Limitations

Despite this being a rapid review of the evidence, we conducted a comprehensive search strategy across multiple databases, with screening conducted by two independent reviewers. The review includes 16 studies providing a depth of evidence from across the globe, supporting the generalisability of findings. We have described these studies in detail, and demonstrated clear themes in the analysis to enhance understanding of the existing evidence. However, as with all rapid reviews, several limitations must be considered. We cannot rule out the possibility that other potentially relevant evidence exists which has not been included. Additionally, a number of the included studies were rated moderate or low quality, this was often due to suboptimal reporting of included participants and research methods.

## Conclusion

Findings from this review suggest that in the context of the COVID pandemic, governments seeking to increase the proportion of symptomatic people accessing testing services should:

1. Ensure information on symptoms is clear; that reporting a new, continuous cough, a fever, or a loss of sense of taste or smell is important. It must be emphasised even mild or minor symptoms must be reported without delay;
2. Make testing facilities easy to access and remove any economic barriers associated with their use;
3. Provide assurance that testing facilities are safe from infection;
4. Target the male population to increase their uptake of services.

## Data Availability

Those interested in the data extracted from the included studies may contact the corresponding author

## Appendix A

Ovid MEDLINE(R) and Epub Ahead of Print, In-Process & Other Non-Indexed Citations and Daily 1946 to June 09, 2020

Date searched: 10^th^ June 2020

| # | Search terms | Hits |
| --- | --- | --- |
| 1 | Pandemics/ | 9950 |
| 2 | Coronavirus Infections/ | 9674 |
| 3 | Disease outbreaks/ | 79399 |
| 4 | "avian influenza".mp. | 10791 |
| 5 | Influenza A Virus, H1N1 Subtype/ or "bird flu".mp. or Influenza A Virus, H5N1 Subtype/ | 20603 |
| 6 | Coronavirus/ or coronavirus.mp. or covid 19.mp. or covid-19.mp. | 34377 |
| 7 | Ebola.mp. | 9045 |
| 8 | "equine influenza".mp. | 787 |
| 9 | H1N1.mp. | 21000 |
| 10 | H5N1.mp. | 7902 |
| 11 | MERs.mp. or Middle East Respiratory Syndrome Coronavirus/ | 4868 |
| 12 | "Middle East respiratory syndrome".mp. | 2325 |
| 13 | pandemic*.mp. | 37034 |
| 14 | Epidemics/ or epidemic*.mp. | 109445 |
| 15 | SARS Virus/ or SARs.mp. or Severe Acute Respiratory Syndrome/ | 16796 |
| 16 | "severe acute respiratory syndrome".mp. | 12771 |
| 17 | "swine flu".mp. | 956 |
| 18 | Help-Seeking Behavior/ | 745 |
| 19 | Help-seek*.mp. | 6215 |
| 20 | (symptom* adj3 report*).mp. | 41088 |
| 21 | (request* or ask* or demand* or call or contact* or access* or inquire* or enquire* or consult* or seek* or requir* or appl* or order* or need* or want*) adj3 (test*) | 94989 |
| 22 | (test* or diagnos*) adj3 (stigma* or fear* or worr* or shame* or reluctan* or knowledge or recogni* or risk perception or perceived risk) | 26496 |
| 23 | Or/1-17 | 248141 |
| 24 | Or/18-22 | 167232 |
| 25 | 23 and 24 | 2169 |
| 26 | limit 25 to humans | 1542 |
| 27 | limit 26 to english language | 1418 |

APA PsycInfo 1806 to June Week 1 2020

Date searched: 10^th^ June 2020

APA PsycInfo 1806 to June Week 1 2020 Date searched: 10<sup>th</sup> June 2020
| # | Search terms | Hits |
| --- | --- | --- |
| 1 | exp Pandemics/ | 534 |
| 2 | "avian influenza".mp. | 114 |
| 3 | "bird flu".mp. | 34 |
| 4 | coronavirus.mp. or covid 19.mp. or covid-19.mp. | 280 |
| 5 | Ebola.mp. | 414 |
| 6 | "equine influenza".mp. | 0 |
| 7 | H1N1.mp. | 495 |
| 8 | H5N1.mp. | 57 |
| 9 | MERs.mp. or "Middle East respiratory syndrome".mp. | 99 |
| 10 | pandemic*.mp. | 2015 |
| 11 | epidemic.mp. or exp Epidemics/ | 12570 |
| 12 | SARs.mp. or "severe acute respiratory syndrome".mp. | 525 |
| 13 | "swine flu".mp. | 92 |
| 14 | exp Help Seeking Behavior/ | 13495 |
| 15 | Help-seek*.mp. | 10538 |
| 16 | (symptom* adj3 report*).mp. | 19817 |
| 17 | (request* or ask* or demand* or call or contact* or access* or inquire* or enquire* or consult* or seek* or requir* or appl* or order* or need* or want*) adj3 (test*) | 25925 |
| 18 | (test* or diagnos*) adj3 (stigma* or fear* or worr* or shame* or reluctan* or knowledge or recogni* or risk perception or perceived risk) | 22000 |
| 19 | Or/1-13 | 14733 |
| 20 | Or/14-18 | 84131 |
| 21 | 19 and 20 | 267 |
| 22 | limit 21 to humans | 257 |
| 23 | limit 22 to english language | 253 |

ProQuest (Coronavirus Research Database, Public Health Database, Social Science Database, Sociology Database and Internal Bibliography of the Social Science [IBSS])

Date searched: 11^th^ June 2020

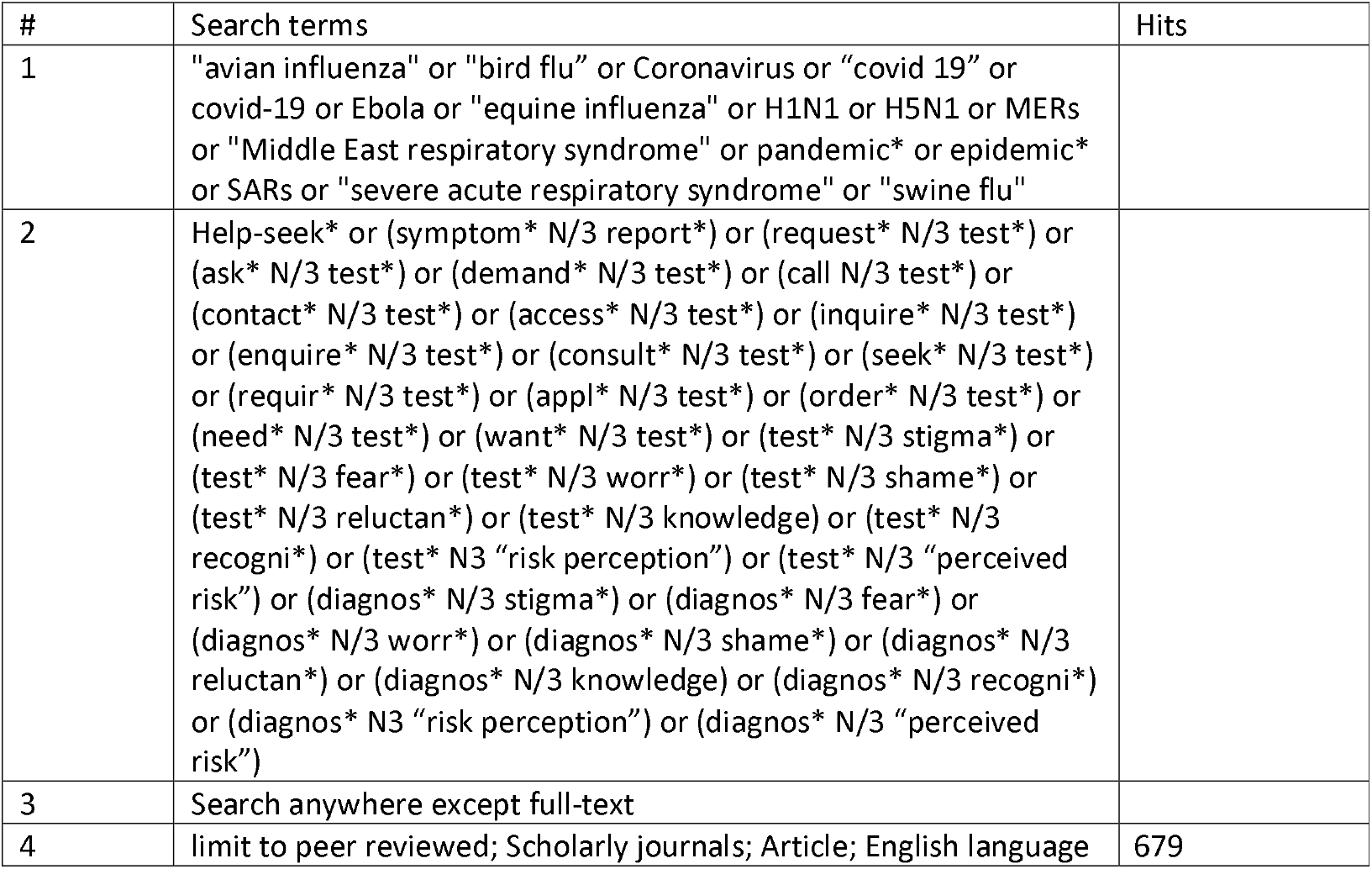

| # | Search terms | Hits |
| --- | --- | --- |
| 1 | "avian influenza" or "bird flu" or Coronavirus or "covid 19" or covid-19 or Ebola or "equine influenza" or H1N1 or H5N1 or MERs or "Middle East respiratory syndrome" or pandemic* or epidemic* or SARs or "severe acute respiratory syndrome" or "swine flu" |  |
| 2 | Help-seek* or (symptom* N/3 report*) or (request* N/3 test*) or (ask* N/3 test*) or (demand* N/3 test*) or (call N/3 test*) or (contact* N/3 test*) or (access* N/3 test*) or (inquire* N/3 test*) or (enquire* N/3 test*) or (consult* N/3 test*) or (seek* N/3 test*) or (requir* N/3 test*) or (appl* N/3 test*) or (order* N/3 test*) or (need* N/3 test*) or (want* N/3 test*) or (test* N/3 stigma*) or (test* N/3 fear*) or (test* N/3 worr*) or (test* N/3 shame*) or (test* N/3 reluctan*) or (test* N/3 knowledge) or (test* N/3 recogni*) or (test* N3 "risk perception") or (test* N/3 "perceived risk") or (diagnos* N/3 stigma*) or (diagnos* N/3 fear*) or (diagnos* N/3 worr*) or (diagnos* N/3 shame*) or (diagnos* N/3 reluctan*) or (diagnos* N/3 knowledge) or (diagnos* N/3 recogni*) or (diagnos* N3 "risk perception") or (diagnos* N/3 "perceived risk") |  |
| 3 | Search anywhere except full-text |  |
| 4 | limit to peer reviewed; Scholarly journals; Article; English language | 679 |

## Appendix B

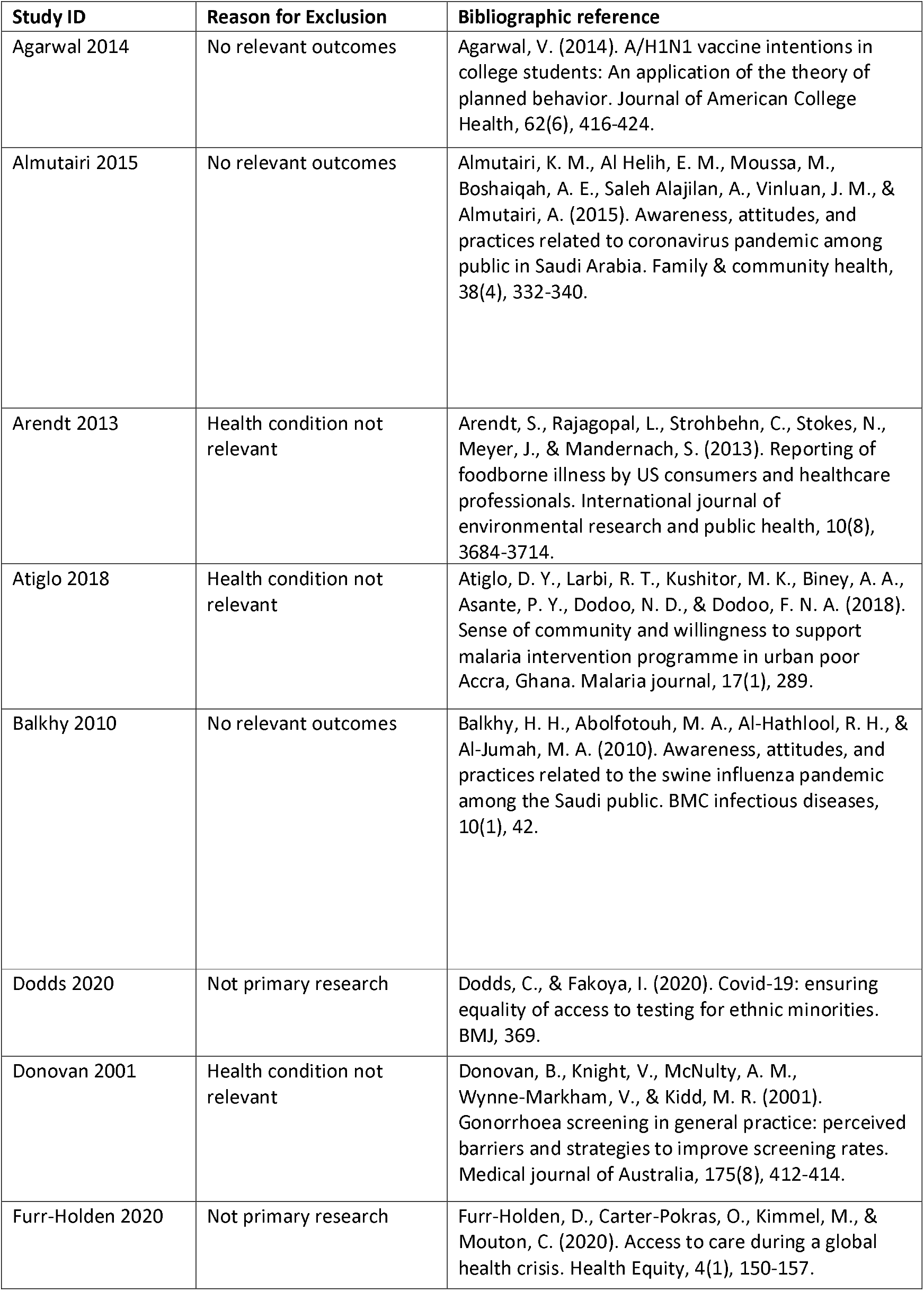
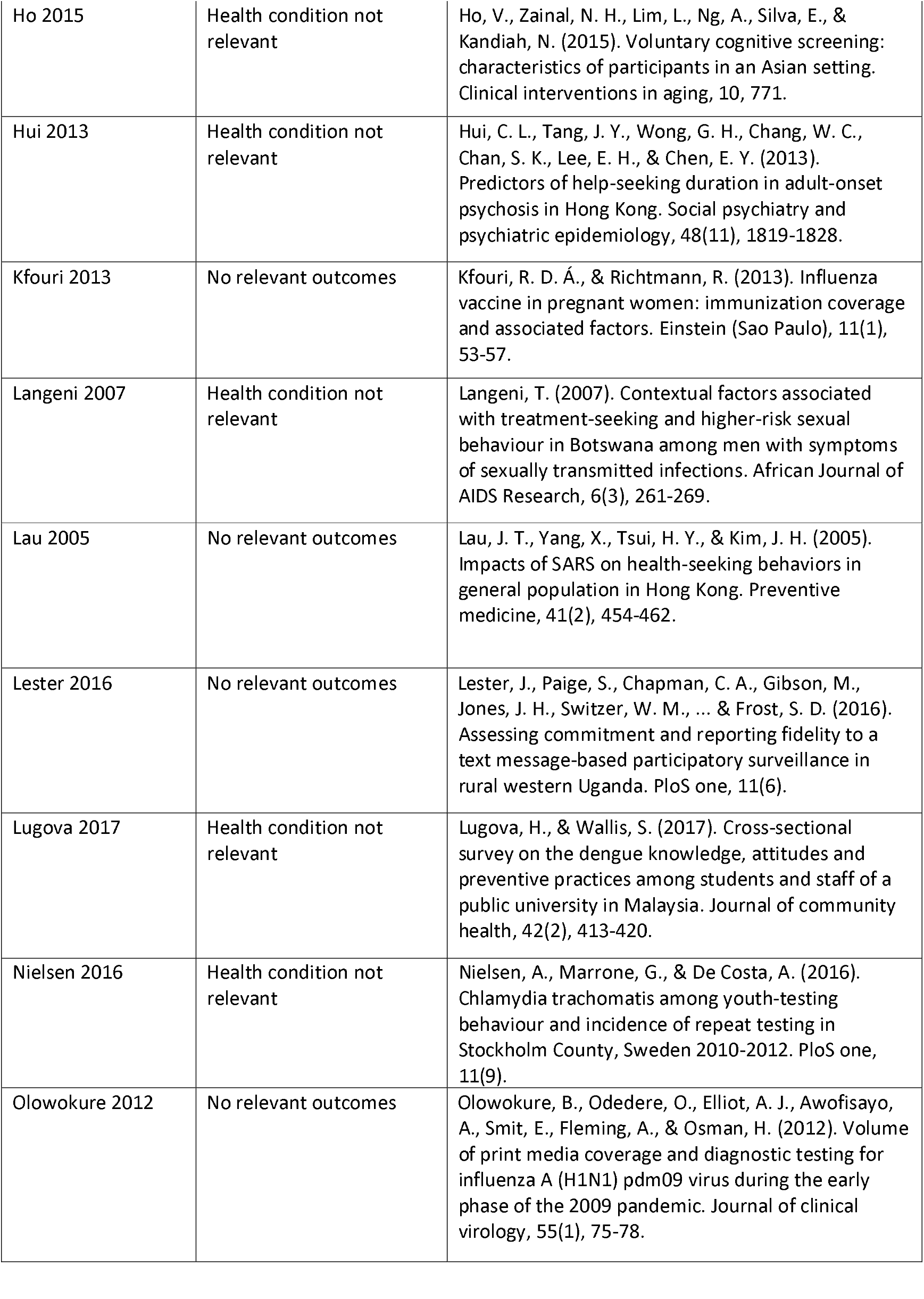
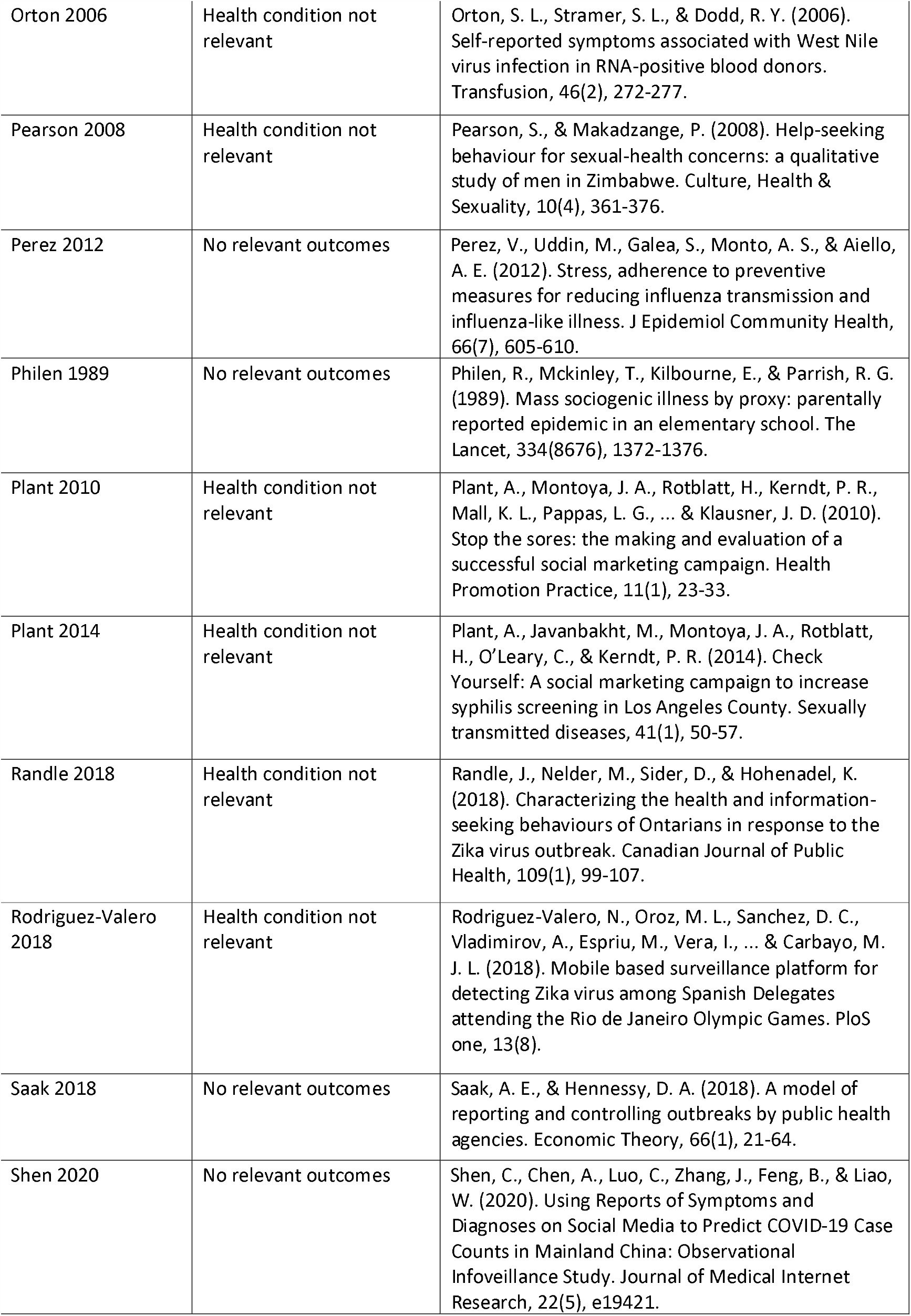
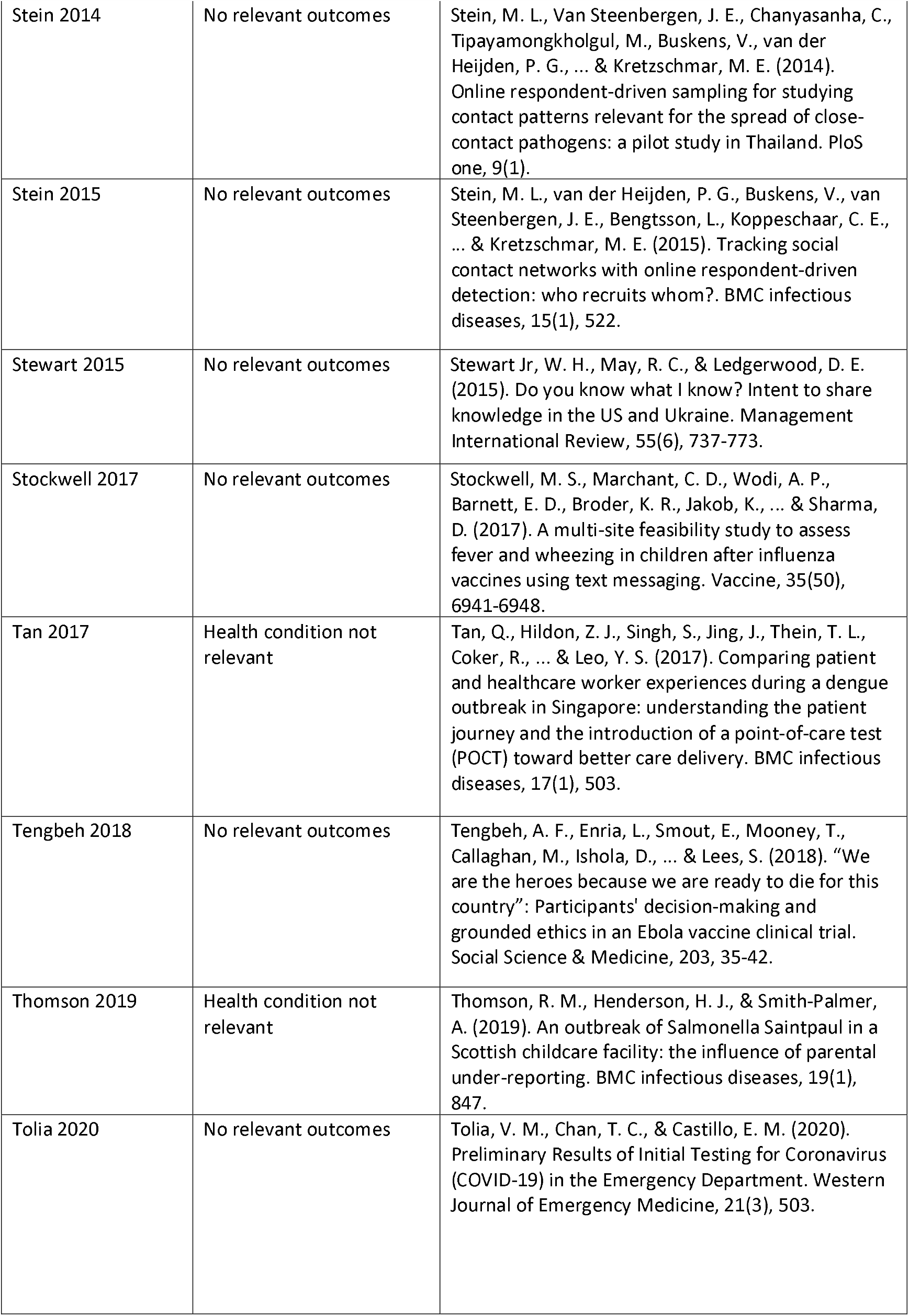
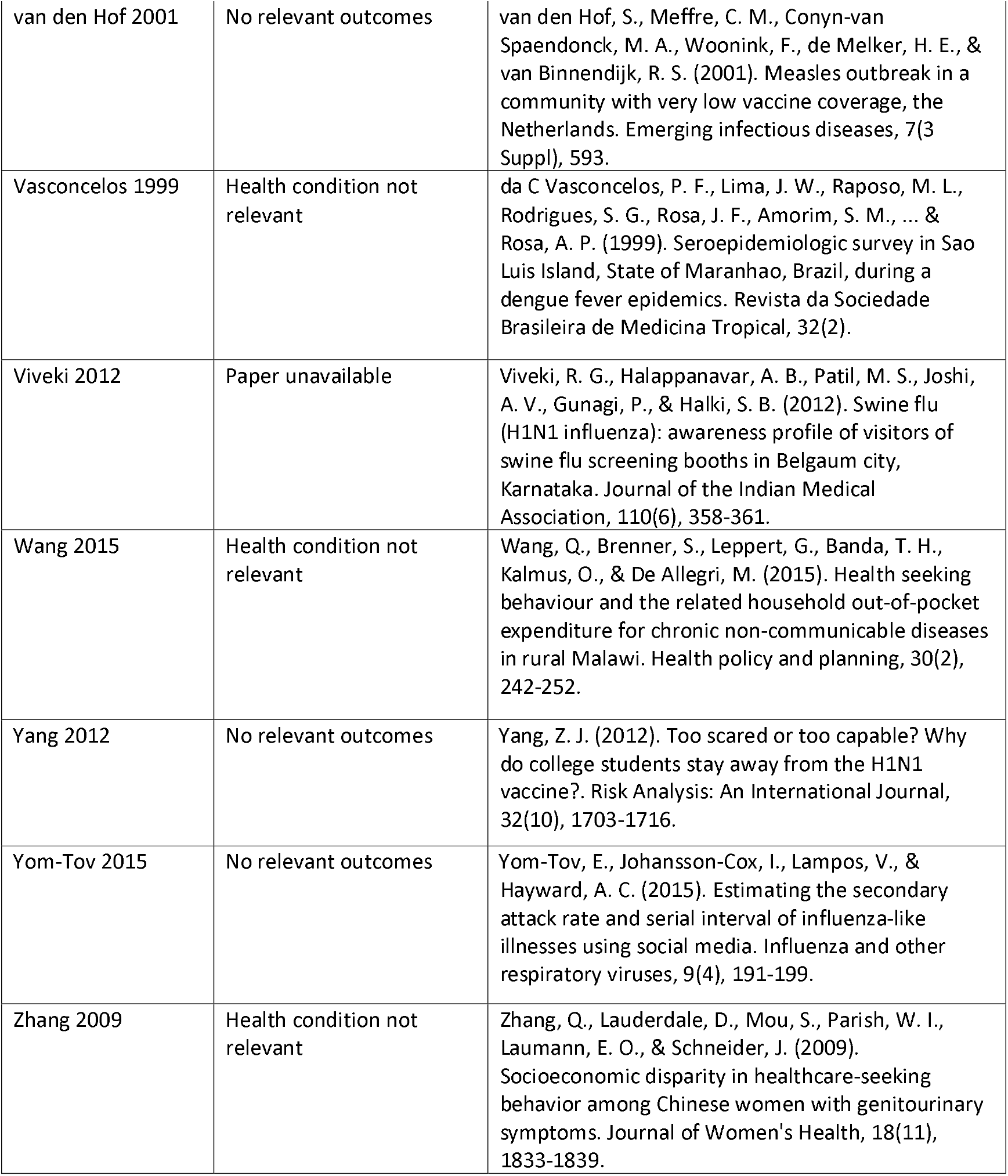

